# Mental Health of Clinical Staff Working in High-Risk Epidemic and Pandemic Health Emergencies: A Rapid Review of the Evidence and Meta-Analysis

**DOI:** 10.1101/2020.04.28.20082669

**Authors:** Vaughan Bell, Dorothy Wade

## Abstract

The global pandemic of SARS-CoV-2 / COVID-19 has raised concerns about the potential mental health impact on frontline clinical staff. However, given that poor mental health is common in staff working in acute medicine, we aimed to estimate the additional burden of working directly with infected patients during epidemic and pandemic health emergencies. We completed a rapid review of the evidence and identified 74 relevant studies from outbreaks of COVID-19, Ebola, H1N1 influenza, Middle East respiratory syndrome (MERS), and severe acute respiratory syndrome (SARS). Due to varying caseness criteria, a meta-analysis of prevalence was not possible. However, it was clear that levels of self-reported depression, anxiety and posttraumatic stress disorder (PTSD) related symptoms were high, and somewhat higher in clinical staff working in high exposure roles. To assess the impact of high- versus low-exposure healthcare work more formally, we estimated the standardised mean difference (SMD) of scale means using a random effects meta-analysis. High exposure work was associated with only a small additional burden of acute mental health problems compared to low exposure work (anxiety: SMD=0.22, 95% CI 0.06 – 0.38; PTSD symptoms: SMD=0.21, 95% CI 0.01 – 0.4; depression: SMD=0.20, -0.07 – 0.47). This effect was potentially inflated by publication bias and there was a moderate risk of bias in the studies in the meta-analysis. A narrative review of candidate risk factors identified being a nurse, seeing colleagues infected, experiencing quarantine, non-voluntary role assignment, and experiencing stigma, as associated with particularly poor mental health outcomes. Protective factors included team and institutional support, use and faith in infection prevention measures, and a sense of professional duty and altruistic acceptance of risk. Notably, formal psychological support services were valued by frontline staff, although those with the highest burden of mental health difficulties were the least likely to request or receive support.

## Introduction

The recent SARS-CoV-2 / COVID-19 pandemic has seen an increased demand on clinical staff, who need to treat large numbers of patients, often in newly-purposed wards, with little disease-specific evidence to guide treatment. Many clinical staff have been moved into new roles and may be managing acutely unwell patients using unfamiliar equipment. Stresses caused by high patient mortality rates, staffing shortages, concerns about infecting self or family members, and changing guidance on personal protective equipment can add to work pressure. This has raised concerns about the potential impact on the mental health of epidemic and pandemic responders (Chen et al., 2020; Greenberg et al., 2020).

However, high rates of poor mental health are common in clinical staff working in acute medicine generally (Carrieri et al., 2018; He et al., 2020; Su et al., 2009) and so estimating the additional impact of epidemic and pandemic response, not solely the extent of poor mental health, is also important in guiding decisions to protect staff’s mental health during times of increased demand on clinical services. In doing so, identifying risk and protective factors for mental health outcomes is key.

Consequently, we review and meta-analyse studies on the mental health of clinical staff dealing with epidemics and pandemics of high-risk infectious diseases, including studies from coronavirus disease 2019 (COVID-19), Ebola virus disease, H1N1 influenza, severe acute respiratory syndrome (SARS), and middle east respiratory syndrome (MERS) to understand the potential impact on mental health and to inform policy on supporting staff during the current COVID-19 pandemic.

## Methods

There is no standardised procedure for conducting rapid reviews, although several approaches have been used to reduce the complexity of the review process (Haby et al., 2016). We used an iterative rapid review procedure that selected search terms with a high sensitivity for relevant articles and then used the reference lists to identify further articles. We searched PubMed, Medline, PsychInfo and Embase for articles including ‘mental health’ or ‘psychosocial’ or ‘emotional’ and ‘staff’ and a number of disease specific key words (epidemic, epidemics, pandemic, flu, SARS, MERS, COVID-19, Ebola, Marburg, H1N1, H7N6) in the title. We also searched pre-print servers MedRXviv and SSRN for pre-prints relating to COVID-19.

**Figure 1.**
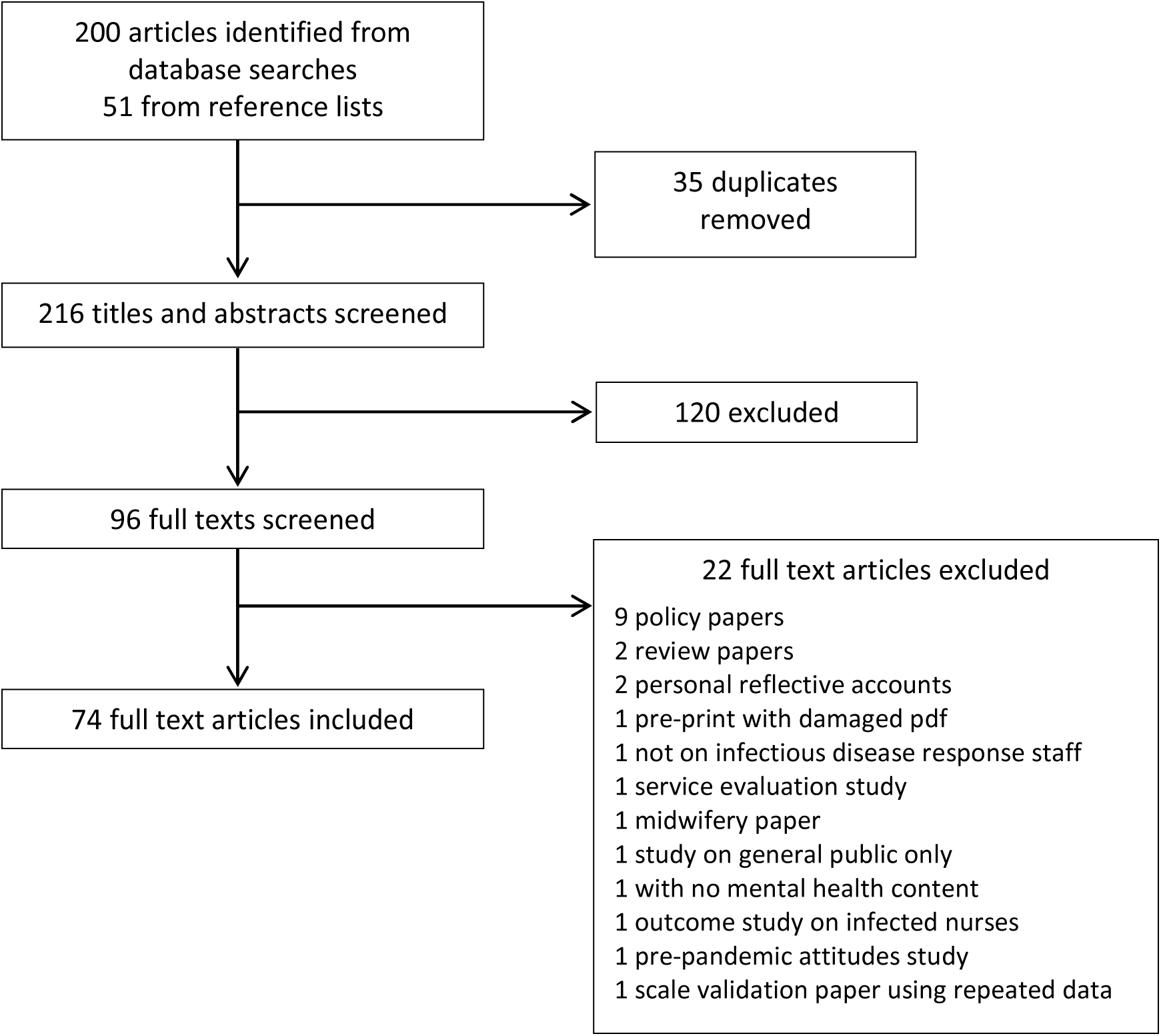
Study screening process

Duplicate articles were removed and titles and abstracts were screened for relevance. The full text of the remaining articles was read. Reference lists were searched for additional relevant articles and these were reviewed for inclusion using the same method as articles found through database searches. Articles were included if they were peer-reviewed articles published in English or Spanish (the languages of the authors) that reported mental health outcomes in clinical staff managing high-risk infectious disease outbreaks. One article in Chinese with numeric results reported in the English-language abstract was also included. Studies of all methodologies reporting original results were included (including qualitative and quantitative studies).

The final count of articles included by condition were: COVID-19: 14, Ebola: 11, H1N1: 4, MERS: 6, SARS: 39. No relevant studies on Marburg or H7N6 flu were found. Studies were conducted in China (16), Hong Kong (12), Taiwan (11), Canada (8), Singapore (6), Sierra Leone (4), Saudi Arabia (3), South Korea (3), Germany (1), Greece (1), Israel (1), Japan (1), Liberia (1), Mexico (1), Uganda and the Republic of Congo (1), United States (2), and two studies that recruited aid workers who had worked in various West African countries.

Numeric data from studies reporting i) above-cut off prevalence or, ii) means and standard deviations from validated anxiety, posttraumatic stress disorder (PTSD) and depression symptom scales were collated. Due to studies using differing cut-offs for determining caseness even when using the same scale, meta-analysis for prevalence was not possible and so a narrative review of prevalence was conducted. However, meta-analysis for differences in mean scale scores between high and low exposure roles was possible and was used to help determine the mental health impact of work involving high exposure to infected patients in epidemic and pandemic health emergencies. We defined high and low exposure as direct contact with infected patients, or work in wards where direct contact was considered highly likely (e.g. critical care, designated treatment wards, emergency departments) compared to clinicians working without direct contact or work in hospital areas where direct contact was unlikely. A narrative review of risk factors was conducted across all studies regardless of methodology.

We conducted a power analysis for meta-analysis (Valentine et al., 2010) to determine the minimum number of studies required to detect a statistically significant difference in standardised mean difference between high and low exposure groups. We calculated power to detect a small effect size and based on results of the Brooks et al. (2018) systematic review of SARS response studies, assumed medium study heterogeneity and an average group size of N=150. This indicated that a minimum study count of 5 was needed. Effect sizes were calculated as standardised mean differences and we used a random effects model to estimate pooled effect sizes. The possibility of publication bias was assessed using trim and fill (Duval and Tweedie, 2000), rank correlation test of funnel plot asymmetry (Begg and Mazumdar, 1994), and Egger’s test (Egger and Smith, 1998). All analysis was conducted with R (version 3.6.1) using the ‘meta’ package (version 4.11.0) and was conducted on a Linux x86_64 platform.

A risk of bias assessment was conducted for all studies reporting prevalences or included in the meta-analysis.

Following recommendations for research responding to the COVID-19 pandemic (Holmes et al., 2020) all data, and Jupyter notebooks including analysis code and output, used in this study have been made freely available at the Open Science Framework archive: https://osf.io/tkeh2

## Results

*Prevalence of poor mental health of epidemic and pandemic response healthcare workers* All studies except one (Lancee et al., 2008) used self-report scales to measure symptoms. As can be seen from Tables 1–3, there was considerable variation in the cut-offs used to determine caseness with lower cut-offs understandably producing higher prevalence estimates. Because of this, a meta-analysis of prevalence was not possible.

However, a narrative analysis indicates that the majority of healthcare workers involved in the treatment of patients during high-risk epidemics and pandemics do not report mental health problems, and those that do typically report symptoms in the mild to moderate range with a smaller percentage of responders reporting severe psychopathology. There were only two studies in which the majority of healthcare workers reported symptoms of poor mental health on any measure (COVID-19: Lai et al., 2020; MERS: Lee et al., 2018), and one of these (Lai et al., 2020) used a particularly low cut-off for reporting prevalence (mild range and above).

Although direct statistical comparison between studies is prevented by the use of different cut-off criteria, inspection of the differences in prevalence rates for high and low exposure groups within studies show that high exposure working is associated with higher rates of above cut-off scoring across all measures (mean 29.86%, SD=18.83%) compared to low exposure working (mean 22.82%, SD = 17.01%) with an average additional prevalence of 7.57% in high exposure groups. However, because these are means of above cut-off prevalences where studies have used different cut-offs it is not clear what an additional 7.57% of prevalence represents in terms of severity.

**Table 1.** Above cut-off anxiety symptom prevalence in studies of healthcare responders working in high-risk epidemic and pandemic health emergencies

| *Study* | *Condition* | *Measure* | *Cut-off* | *Sample* | *Prevalence* |  |  | *Timing* |
| --- | --- | --- | --- | --- | --- | --- | --- | --- |
|  |  |  |  |  | *Overall* | *High exposure* | *Low exposure* |  |
| C. Liu et al 2020 | COVID-19 | ZSRAS | >59* | HE and LE | 12.5% | Not given | Not given | During |
| Z. Liu et al 2020 | COVID-19 | ZSRAS | >49 | HE and LE | 16.0% | 18.6% | 14.0% | During |
| J. Z. Huang et al 2020 | COVID-19 | ZSRAS | Not given | HE only | Not relevant | 23.04% | Not relevant | During |
| Lai et al 2020 | COVID-19 | GAD-7 | >4 | HE and LE | 44.6% | 51.3% | 39.4% | During |
| Zhu et al 2020 | COVID-19 | GAD-7 | >7 | HE and LE | 24.36% | 23.66% | 24.83% | During |

HE = High Exposure; LE = Low Exposure; ZSRAS = Zung Self-rating Anxiety Scale; GAD-7 = General Anxiety Disorder-7 scale.

**Table 2.** Above cut-off PTSD symptom prevalence in studies of healthcare responders working in high-risk epidemic and pandemic health emergencies

| *Study* | *Condition* | *Measure* | *Cut-off used* | *Sample* | *Prevalence* |  |  | *Timing* |
| --- | --- | --- | --- | --- | --- | --- | --- | --- |
|  |  |  |  |  | *Overall* | *High exposure* | *Low exposure* |  |
| J. Z. Huang et al 2020 | COVID-19 | PTSD-SS | Not given | HE only | Not relevant | 27.39% | Not relevant | During |
| Lai et al 2020 | COVID-19 | IES-R | >8 | HE and LE | 71.5% | 76.0% | 68.0% | During |
| Zhu et al 2020 | COVID-19 | IES-R | >33 | HE and LE | 30.56% | 33.43% | 28.65% | During |
| Lee et al 2018 | MERS | IES-R | >24 | HE and LE | 51.5% | Not given | Not given | During |
| Chan & Huak 2004 (doctors) | SARS | IES | >29 | HE and LE | Not given | 18.8% | 19.8% | During |
| Chan & Huak 2004 (nurses) | SARS | IES | >29 | HE and LE | Not given | 19.4% | 19.5% | During |
| Chen et al 2005 | SARS | IES-R | >36 | HE and LE | 11% | 17% | 2% | During |
| Su et al 2007 | SARS | DTS-C | >22 | HE and LE | 28.43% | 32.86% | 18.75% | During |
| Lin et al 2007 | SARS | DTS-C | >40 | HE and LE | 19.3% | 21.7% | 13.0% | During |
| Sim et al 2004 | SARS | IES | Idiosyncratic | HE and LE | 9.4% | 7.2% | 10.6% | During |
| Tham et al 2004 | SARS | IES | >25 | Undifferentiated | 17.7% | Not relevant | Not relevant | 6 months after |
| Maunder et al 2006 | SARS | IES | >25 | HE and LE | Not given | 13.8% | 8.4% | 1-2 years after |
| Lancee et al 2008 | SARS | CAPS | DX | HE only | Not relevant | 2% | Not relevant | 1-2 years after |
| Wu et al 2008 | SARS | IES-R | >19 | HE and LE | 10.1% | Not given | Not given | 3 years after |

HE = High Exposure; LE = Low Exposure; PTSD-SS = PTSD Self Report Scale; IES = Impact of Events Scale; IES-R = Impact of Events Scale – Revised; SASR = Stanford Acute Stress Reaction; CIES-R = Chinese Impact of Event Scale-Revised; CAPS = Clinician Administered PTSD Scale; DX = diagnosis.

**Table 3.** Above cut-off depression symptom prevalence in studies of healthcare responders working in high-risk epidemic and pandemic health emergencies

| *Study* | *Condition* | *Measure* | *Cut-off used* | *Sample* | *Prevalence* |  |  | *Timing* |
| --- | --- | --- | --- | --- | --- | --- | --- | --- |
|  |  |  |  |  | *Overall* | *High exposure* | *Low exposure* |  |
| Zhu et al 2020 | COVID-19 | PHQ-9 | >9 | HE and LE | 13.55% | 13.73% | 13.43% | During |
| Lai et al 2020 | COVID-19 | PHQ-9 | >4 | HE and LE | 50.4% | 58.4% | 44.6% | During |
| Chung & Yeung, 2020 | COVID-19 | PHQ-9 | >9 | Undifferentiated | 34.8% | Not given | Not given | During |
| Z. Liu et al 2020 | COVID-19 | ZSRAD | >49 | HE and LE | 34.6% | 37.2% | 32.5% | During |
| Su et al 2007 | SARS | BDI | >9 | HE and LE | 27.45% | 38.57% | 3.13% | During |
| Wu et al 2008 | SARS | CES-D | >15 | HE and LE | 22.8% | Not given | Not given | 3 years after |

HE = High Exposure; LE = Low Exposure; PHQ-9 = Patient Health Questionnaire 9; ZSRAD = Zung Self-rating Depression Scale; BDI = Beck Depression Inventory; CES-D = Center for Epidemiologic Studies Depression Scale

### Meta-analytic estimate of the effect of exposure on acute mental health outcomes

Given that high rates of poor mental health are common in staff working in acute medicine (Carrieri et al., 2018; He et al., 2020; Su et al., 2009) we attempted to estimate the additional impact of working in high exposure roles. Meta-analytic estimates of impact of working in high exposure roles are presented in Figure 2 (anxiety symptom measures), Figure 3 (PTSD symptom measures) and Figure 4 (depression symptom measures).

**Figure 2.**
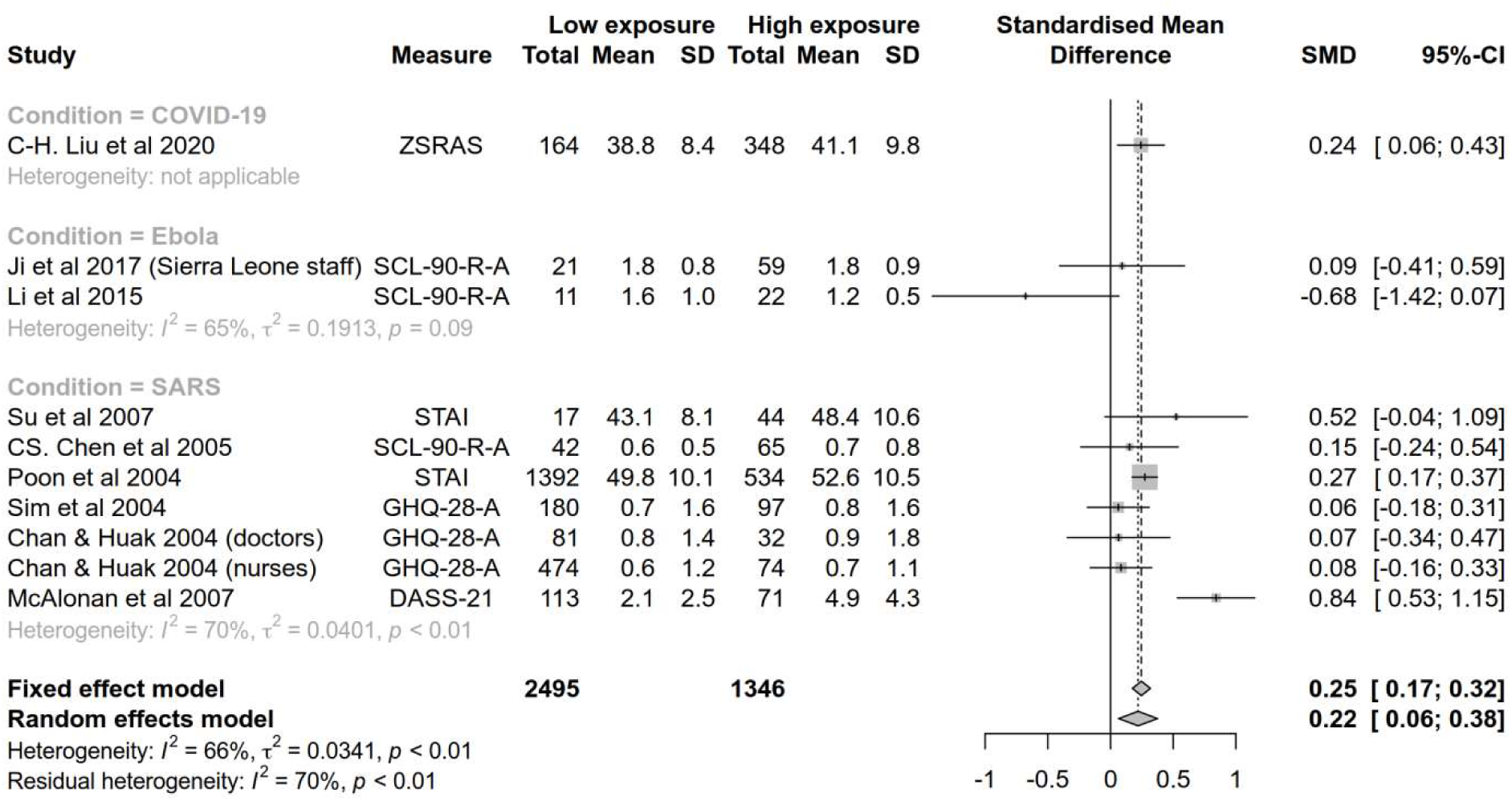
Standardised mean differences in anxiety measures between health care workers with high and low levels of exposure to high-risk epidemic and pandemic diseases

**Figure 3.**
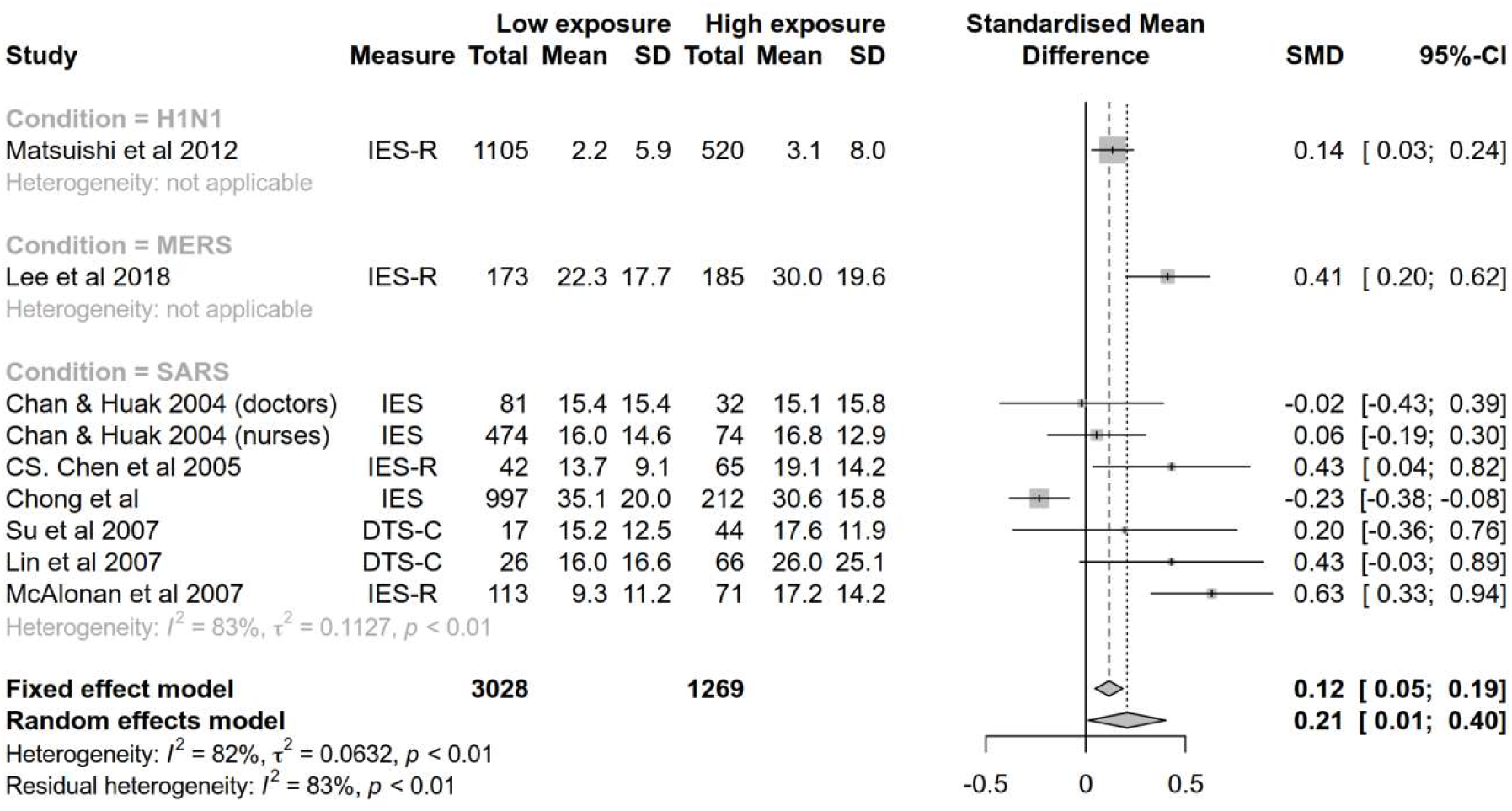
Standardised mean differences in PTSD symptom measures between health care workers with high and low levels of exposure to high-risk epidemic and pandemic diseases

**Figure 4.**
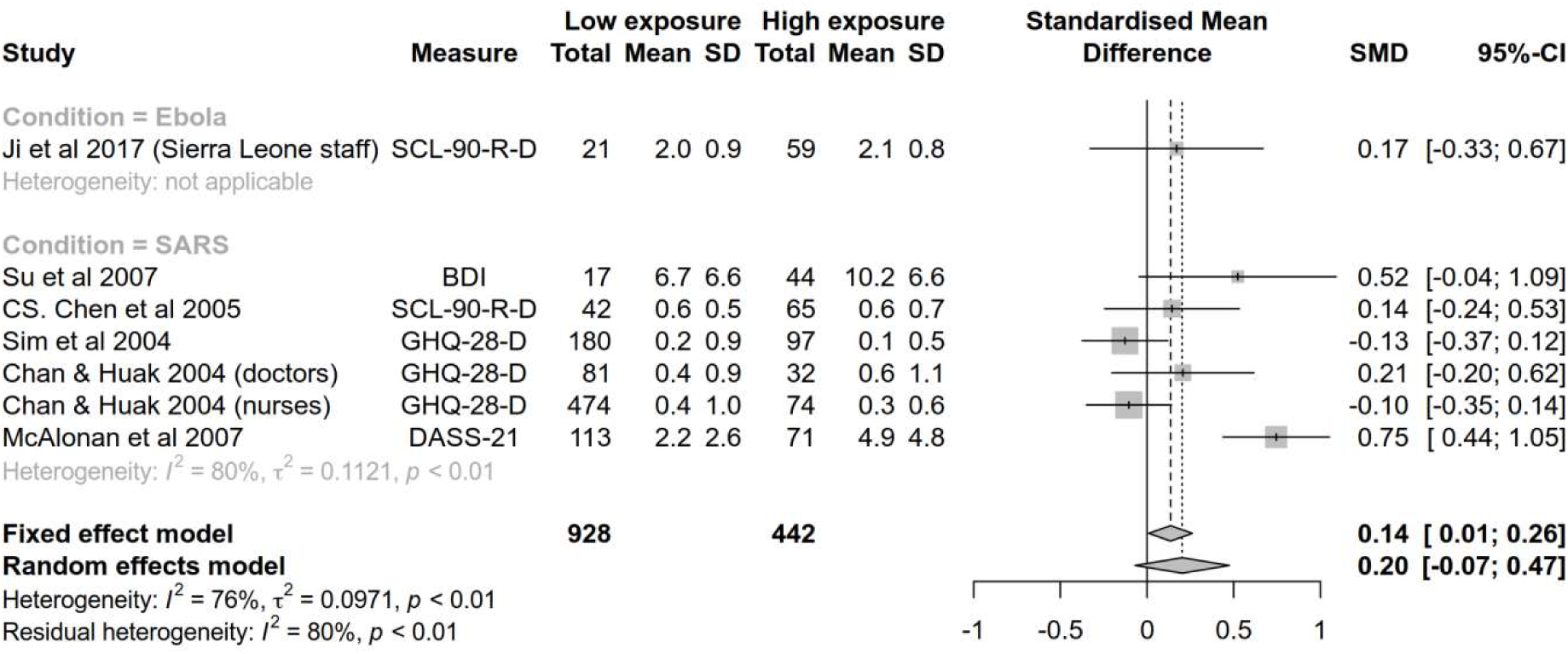
Standardised mean differences in depression measures between health care workers with high and low levels of exposure to high-risk epidemic and pandemic diseases

All analyses show that the difference between working in high and low exposure roles was small (effect sizes of 0.22 and below). The confidence intervals for estimates did not cross zero for anxiety and PTSD symptom measures, but they did cross zero in the case of the meta-analysis for depression measures. However, study heterogeneity was larger than we assumed during the power calculation and there were the fewest number of studies included for the depression meta-analysis (k=6), meaning it was likely under-powered to detect small effects.

Inspection of the funnel plot suggested evidence of publication bias but the statistical tests of funnel plot asymmetry were not interpreted as the minimum criteria of 10 included studies was not met (Ioannidis and Trikalinos, 2007).

### Risk and protective factors

Although it was not possible to examine risk and protective factors meta-analytically, a narrative review was conducted to identify likely candidates.

Nurses typically reported higher levels of symptoms and distress than doctors (Goulia et al., 2010; Huang et al., 2020; Lai et al., 2020; C. Liu et al., 2020; Matsuishi et al., 2012; Nickell et al., 2004; Poon et al., 2004; Tham et al., 2004; Zhu et al., 2020) with a few studies reporting no difference (Dai et al., 2020; Maunder et al., 2006; Sun et al., 2020) and one study reporting higher rates in doctors (Chan and Huak, 2004).

Several studies noted that seeing colleagues infected was a particular source of distress (Dai et al., 2020; Khalid et al., 2016; Raven et al., 2018). Furthermore, being quarantined after infection was reported as a predictor of psychological distress and poor mental health (Bai et al., 2004; Dai et al., 2020; Fiksenbaum et al., 2006; Liu et al., 2012; Marjanovic et al., 2007; Styra et al., 2008; Wu et al., 2009, 2008) which was also reflected in one qualitative study (Robertson et al., 2004) although three studies found no negative impact of quarantine (Chong et al., 2004; Lee et al., 2018; Styra et al., 2008).

Notably, numerous studies reported that clinical staff dealing with high-risk infectious disease experienced stigma from friends, family and the public (Bai et al., 2004; Goulia et al., 2010; Grace et al., 2005; Hewlett and Hewlett, 2005; Khee et al., 2004; Koh et al., 2005; Maunder et al., 2006; McMahon et al., 2016; Meyer et al., 2018; Nickell et al., 2004; Park et al., 2018; Poon et al., 2004; Smith et al., 2017; Styra et al., 2008; von Strauss et al., 2017) and perceived stigma was found to be a predictor of poor mental health in the three studies that looked at this association statistically (Koh et al., 2005; Maunder et al., 2006; Park et al., 2018).

Three studies found that clinical staff who were conscripted, were not willing, or had not volunteered for high exposure roles reported particularly poor mental health outcomes (Chen et al., 2005, 2006; Dai et al., 2020; Tam et al., 2004).

Although small in number, longitudinal studies tended to suggest that symptoms of poor mental health tend to peak early during outbreaks but resolve for the majority of responders as time goes on (Chen et al., 2006; Su et al., 2009). This pattern of initially high levels of anxiety and distress that reduce over time was also reflected in some of the qualitative studies (Chung et al., 2005; Shih et al., 2009). One cross-sectional study conducted at two time points and sampling from the same clinical teams found high levels of self-reported poor mental health for high exposure workers one year after high-exposure work (Maunder et al., 2006) although a study on a subsample of participants using a structured interview assessment found the incidence of new onset mental health problems was essentially no different to that found in the general population (Lancee et al., 2008).

In terms of protective factors that reduced the chance of poor mental health or psychological distress, social support, team cohesion or organisational support were identified by numerous studies (Chan and Huak, 2004; Chang et al., 2006; Fiksenbaum et al., 2006; Khalid et al., 2016; Lee et al., 2005; Marjanovic et al., 2007; Sun et al., 2020; von Strauss et al., 2017; Xiao et al., 2020; Zhu et al., 2020). This theme was reflected in several qualitative studies (Meyer et al., 2018; Raven et al., 2018).

Furthermore, the use, availability, training with, and faith in, infection prevention measures were identified as reducing distress (Chua et al., 2004; Khalid et al., 2016; Lee et al., 2005, 2005; Marjanovic et al., 2007; Maunder et al., 2006; Nickell et al., 2004; Sim et al., 2004; Zhu et al., 2020)

A sense of professional duty and altruistic acceptance of risk were found to be a protective factor in several studies (Goulia et al., 2010; Liu et al., 2012; Oh et al., 2017; Wu et al., 2009) which was a theme that was strongly reflected in qualitative studies (Chung et al., 2005; Hewlett and Hewlett, 2005; Raven et al., 2018; Wong et al., 2012) although one study found accepting the risk of SARS infection as part of the job was not associated with reduced psychopathology (Koh et al., 2005).

Notably, all studies that asked about positive aspects of working in epidemic and pandemic response reported that participants described several factors related to learning and development as an individual and a team (Chua et al., 2004; Grace et al., 2005; Nickell et al., 2004).

### Role of formal psychological support services

Although a recent anecdotal report noted clinicians did not find mental health support particularly useful during COVID-19 response (Chen et al., 2020) several studies found that participants reported formal psychological support services to be a useful source of support (Goulia et al., 2010; Lee et al., 2005; Meyer et al., 2018; Smith et al., 2017; von Strauss et al., 2017). One study specifically asked whether staff needed ‘psychological treatment’ and 8.6% of healthcare workers dealing with COVID-19 reported they did (C. Liu et al., 2020). Conversely, however, Chung and Yeung (2020) reported that only 2% of staff responding to COVID-19 requested psychological support and all “were reassured after a single phone contact by the psychiatric nurse” although this was a notably small study with just 69 participants.

Notably, two large COVID-19 studies suggest that the staff who are most in need of psychological support are the least likely to request or receive it. One study found that high exposure staff were likely to say they needed psychological treatment at half the rate of low exposure staff despite reporting higher levels of psychopathology (C. Liu et al., 2020). In another, clinicians with mental health problems were less likely to receive psychological support than clinicians without (Z. Liu et al., 2020).

## Discussion

To estimate the impact of high exposure work in epidemic and pandemic health emergencies, we completed a rapid review and meta-analysis of studies reporting on the mental health of clinical staff working in high-risk epidemic and pandemic health emergencies, including studies from the recent COVID-19 pandemic. Both of these comparisons suggest that the impact of epidemic and pandemic response work on mental health is small, but in the case of meta-analytic assessment, statistically detectable. However, this is in addition to already high levels of poor mental health that are common in acute medical staff. A narrative review of potential risk factors for poor mental health in epidemic and pandemic response identified being a nurse, experiencing stigma from others, seeing colleagues infected, and being personally quarantined as predictors of worse outcomes. Protective factors included social and occupational support, effectiveness and faith in infection control measures, a sense of professional duty and altruistic acceptance of risk. Formal psychological support services were identified as a valuable form of support.

It is worth noting some of the shortcomings of the evidence used to inform this review. More studies reported results from the analysis of mental health measures than reported sufficient detail (prevalence, mean scores etc) to allow them to be included in the numeric assessment of results in this review. Study quality was moderate at best and there was a suggestion of publication bias. Furthermore, studies almost exclusively used self-report measures rather than structured interview assessments.

The results of this review and meta-analysis raise several issues with regard to provide ongoing and long-term support for clinical staff responding to such health emergencies. One is the extent to which epidemic and pandemic response is uniquely ‘traumatising’ and might lead to high levels of posttraumatic stress disorder, thereby implying that a trauma-focus for staff support should be the dominant approach to addressing outcomes of poor mental health. With regard to the published studies, although there are seemingly high rates of staff who score above cut-off on measures of PTSD symptoms, some significant caveats need to be born in mind. As can be seen from Table 3, cut-off values used for defining a positive ‘case’ varied considerably even between studies using the same measure. The most widely used scale in these studies is the Impact of Events Scale-Revised for which a cut-off of 33–34 has been found to be the most predictive of diagnosable PTSD (Creamer et al., 2003; Morina et al., 2013) and yet most studies use a cut-off of considerably less. This suggests that an important proportion of those reported under the prevalence figures are likely to have transitory, sub-syndromal PTSD symptoms, or non-specific distress, that may be a risk for PTSD but are unlikely to reach the level of a diagnosable case. Indeed, the prevalences reported here are comparable to prevalences found in clinical staff more widely. For example, the reported prevalence of >33 scoring on the IES-R is 15% in acute medical staff (Naumann et al., 2017), 16% in surgical trainees (Thompson et al., 2017), and 17% in cancer physicians (McFarland and Roth, 2017). Studies included in this review using similar IES-R cut-offs tended to report lower prevalence rates with only one study (Zhu et al., 2020) reporting higher.

Furthermore, studies often did not differentiate between PTSD symptoms arising from pandemic and epidemic response work and those from other events meaning it is not clear to what extent epidemic or pandemic response work was the key causal factor. Finally, symptoms were almost exclusively measured by self-report measures which are known to inflate the rate of true cases (Bonanno et al., 2010). Indeed, high rates on self-report measures but low rates on structured interview assessments have been found in SARS responders (Lancee et al., 2008).

This suggests that although there is potential for trauma and this should be included in considerations for staff support, it is currently not clear that PTSD is an outcome particularly associated with epidemic and pandemic response meaning support efforts should be ‘trauma ready’ rather than ‘trauma focused’. However, we note here the importance of contextual factors. The capacity of the healthcare system, and indeed the population, are key factors in determining the impact of the medical response on the staff responsible for delivering it.

Indeed, some of the contextual factors were reflected in the risk and protective factors identified in the review. These chime with previous work on epidemic response work (Brooks et al., 2018b) and the wider literature on mental health outcomes in high-risk work (Brooks et al., 2019) and suggest similar measures to support staff, namely promoting good leadership and team cohesion, maintaining high standards of infection control and training. In addition, this review highlights that additional attention should be given to nurses, those affected by seeing colleagues infected, where staff are quarantined after being infected, and where individuals experience stigma from others.

It is worth highlighting that formal psychological support was considered useful by clinical staff but that it was least requested and less frequently received by those with higher levels of mental health difficulties. Although voluntary engagement with mental health services is considered the ideal model to avoid potential iatrogenic effects (Brooks et al., 2018a), particular attention should be paid to pathways to formal support to make these as accessible as possible for those who need them. This is particularly important as staff working in acute medicine may already have high levels of poor mental health and access to effective treatment during health emergencies should be a priority.

### Priorities for research

The majority of studies are cross-sectional measuring mental health in staff working in epidemic and pandemic response. These studies typically have large sample sizes and good response rates. However, there remains a need for: i) standardisation of measures, reporting, and criteria for prevalence; ii) adequately sampled case-control studies that compare epidemic and pandemic response staff to control groups of other staff in acute medicine; and iii) longitudinal studies to examine the course of psychological distress over time. We also note that i) would effectively be solved if open data was available from the relevant studies, and we strongly encourage researchers to make this data available, particularly when researching infectious disease health emergencies.

## Data Availability

All data and analysis code have been made freely available at the study's Open Science Framework archive.

https://osf.io/tkeh2

## Notes

### Competing Interest Statement

The authors have declared no competing interest.

### Funding Statement

The authors received no external funding for the completion of this study.

